# Multiple sclerosis cortical lesion detection with deep learning at ultra-high-field MRI

**DOI:** 10.1101/2022.03.02.22271269

**Authors:** Francesco La Rosa, Erin S. Beck, Josefina Maranzano, Ramona-Alexandra Todea, Peter van Gelderen, Jacco A. de Zwart, Nicholas J. Luciano, Jeff H. Duyn, Jean-Philippe Thiran, Cristina Granziera, Daniel S. Reich, Pascal Sati, Meritxell Bach Cuadra

## Abstract

Manually segmenting multiple sclerosis (MS) cortical lesions (CL) is extremely time-consuming, and past studies have shown only moderate inter-rater reliability. To accelerate this task, we developed a deep learning-based framework (CLAIMS: Cortical Lesion Artificial Intelligence-based assessment in Multiple Sclerosis) for the automated detection and classification of MS CL with 7T MRI.

Two 7T datasets, acquired at different sites, were considered. The first consisted of 60 scans that include 0.5mm isotropic MP2RAGE acquired 4 times (MP2RAGEx4), 0.7mm MP2RAGE, 0.5mm T2*-weighted GRE, and 0.5mm T2*-weighted EPI. The second dataset consisted of 20 scans including only 0.75×0.75×0.9 mm MP2RAGE. CLAIMS was first evaluated using 6-fold cross-validation with single and multi-contrast 0.5mm MRI input. Second, performance of the model was tested on 0.7mm MP2RAGE images after training with either 0.5mm MP2RAGEx4, 0.7mm MP2RAGE, or alternating the two. Third, its generalizability was evaluated on the second external dataset and compared with a state-of-the-art technique based on partial volume estimation and topological constraints (MSLAST). CLAIMS trained only with MP2RAGEx4 achieved comparable results to the multi-contrast model, reaching a CL true positive rate of 74% with a false positive rate of 30%. Detection rate was excellent for leukocortical and subpial lesions (83%, and 70%, respectively), whereas it reached 53% for intracortical lesions. The correlation between disability measures and CL count was similar for manual and CLAIMS lesion counts. Applying a domain-scanner adaptation approach and testing CLAIMS on the second dataset, the performance was superior to MSLAST when considering a minimum lesion volume of 6μL (lesion-wise detection rate of 71% vs 48%).

The proposed framework outperforms previous state-of-the-art methods for automated CL detection across scanners and protocols. In the future, CLAIMS may be useful to support clinical decisions at 7T MRI, especially in the field of diagnosis and differential diagnosis of multiple sclerosis patients.

## Introduction

Multiple sclerosis (MS) is an inflammatory demyelinating disease affecting the central nervous system. It is characterized by focal areas of white matter (WM) demyelination^1^. In recent decades, however, histopathological studies have shown that lesions in the cortex are also common^1–3^. Moreover, increasing use of ultra-high-field MRI has led to the observation that cortical lesions (CL) are extremely frequent in MS patients, persist over time, correlate with disability and progressive disease^4,5^, and may help to differentiate MS from its clinical mimics. CL have been classified into three major types with potentially different etiologies^1^: leukocortical (type 1, located at the interface between WM and GM), intracortical (type 2, involving purely the cortex and not reaching the pial surface), and subpial (including type 3 lesions located entirely in the GM and touching the pial surface, and type 4 lesions, extending from the pial surface, through the cortex, into the white matter). In order to maximize MS diagnostic and prognostic accuracy, it is, therefore, crucial to analyze the clinical implication of CL and their response to current and novel MS treatments.

Contrary to white matter lesions (WML), CL, and particularly intracortical and subpial lesions, are difficult to visualize with conventional sequences, although in recent years, the development of advanced sequences has led to improved CL visualization. However, at 3T, even advanced sequences such as double inversion recovery (DIR), magnetization-prepared 2 rapid acquisition gradient echoes (MP2RAGE), and phase sensitive inversion recovery (PSIR) are relatively insensitive to CL^6,7^. Compared to lower magnetic fields, ultra-high field MRI allows higher signal-to-noise ratio (SNR) and enhanced magnetic susceptibility contrast, both of which can provide important insights into MS pathophysiology^8^. At 7T, T2* weighted methods and MP2RAGE dramatically improve CL, and especially subpial lesion, visualization^6,7,9–12^. For example, compared to the combined use of 7T MP2RAGE and T2*w images, 3T double inversion recovery was 6% sensitive for subpial lesions and 3T MP2RAGE was 5% sensitive^6^. Thus, 7T imaging is essential for expanding our understanding of cortical demyelination^13^. With the recent FDA approval of 7T scanners for clinical use, we can expect an increasing number of scans on these devices in the coming years. However, even with the most sensitive imaging methods, identification and segmentation of cortical lesions is very time-consuming and requires significant experience.

Despite the promise of 7T for the visualization of CL, studies using 7T MRI have shown only modest inter-rater reliability, both when considering CL detection and CL count, assessed with the Cohen’s kappa coefficient (k) and the Lin’s concordance correlation coefficient (CCC), respectively^3,11^. In Nielsen et al., for instance, two experts analyzing 7T FLASH-T2* images, and a total of 103 CL had a moderate inter-rater agreement (k=0.69)^11^. In a similar study, Harrison et al. considered 7T magnetization-prepared rapid acquisition gradient echo (MPRAGE) and assessed the intra-rater and inter-rater reliability in terms of lesion count by two experts^3^. Strong intra-rater agreement was found (CCC = 0.96), whereas the inter-rater correlation was weak (CCC = 0.54). An automatic segmentation method for CL will therefore be essential to support large-scale studies and consistent evaluation of cortical lesions in multi-site clinical trials.

Deep learning methods have lately shown outstanding performance in MRI segmentation, classification, and synthesis^14^. Applied to MS, these techniques have achieved state-of-the-art performance for WML segmentation^15–17^ as well as for the automated assessment of other novel imaging biomarkers, such as paramagnetic rim lesions and the central vein sign^18,19^. However, while some different approaches have been presented to automatically segment CL based on 3T MRI ^20–22^, to the best of our knowledge, only two methods have been proposed for detection at 7T^23,24^. First, Fartaria et al. have proposed MSLAST (Multiple Sclerosis Lesion Analysis at Seven Tesla), an automated method based on partial volume estimation and topological constraints that segments both WML and CL with a single MP2RAGE scan as input. MSLAST was evaluated on a cohort of 25 individuals with MS imaged in two research centers using 0.7mm isotropic MP2RAGE images and achieved a detection rate of 74% and 58% for WML and CL, respectively, with a false positive rate of 40% when considering a minimum lesion size of 6 μL^23^. Second, we previously proposed a deep learning-based method to detect and classify CL in 7T MRI considering three different contrasts: MP2RAGE, T2*w GRE, and T2*w EPI (voxel size of 0.5×0.5.x0.5 mm). On a larger cohort of 60 MS patients, the performance of our method showed promise, achieving a CL detection rate of 67% and a false positive rate of 42%. Moreover, almost 400 CL (24% of the total false positives) detected by our proposed network and initially classified as false positives, were retrospectively judged as lesions by an expert.

The contribution of the current work is three-fold. First, building upon our previous method^24^, we propose “CLAIMS” (Cortical Lesion AI-based assessment in Multiple Sclerosis), an improved pipeline for the automated detection and classification of CL with either single or multi-contrast MRI at 7T. Second, we assess the relative value of MP2RAGE and T2*-weighted images, as well as the impact of MP2RAGE image quality, for this automated task. Third, an additional dataset from another institution is considered in order to test CLAIMS’s generalizability and robustness. For this purpose, a domain adaptation approach was performed and evaluated as well. Importantly, we evaluated CLAIMS performance with respect to our experts’ MRI-based annotations, which throughout the manuscript are referred to as ground truth.

## Material and Methods

### Datasets

Two datasets, acquired at Institution A and B, were analyzed in this study. At Institution A, MRI acquisitions were performed on 60 individuals with MS (43 relapsing remitting, 17 progressive, 63% female, 49±11 (mean ± standard deviation) years old, age range 29–77 years) with Expanded Disability Status Scale (EDSS) scores ranging from 0 to 7.5 (median 2.0) and disease duration of 14±11 (range 0-42) years. In addition to EDSS, the clinical assessment included the following disability measures: 9-Hole Peg Test (9-HPT), 25-foot timed walk (25TW), and Symbol Digit Modalities Test (SDMT). Imaging was done on a 7T whole-body research system (Siemens Healthcare, Erlangen, Germany) using a 32-channel head coil. The MRI protocol included: (i) 3D MP2RAGE (TR/TI1/TI2/TE = 6000/800/2700/5 ms, voxel size = 0.5×0.5×0.5 mm) repeated 4 (total acquisition time ∼40 minutes), (ii) 3D-segmented T2*w EPI [20, 21] (TR/TE = 52/23 ms, voxel size = 0.5×0.5×0.5 mm) acquired in two partially overlapping volumes for whole brain coverage (total acquisition time ∼7 minutes), (iii) 2D T2*w multi-echo GRE (TR/TE1/TE2/TE3/TE4/TE5 = 4095/11/23/34/45/56 ms, voxel size = 0.215×0.215×1.0 mm) acquired in three volumes for nearly full supratentorial coverage and averaged across the echo times (total acquisition time ∼35 min). Moreover, for 55 out of the 60 patients, an additional single 3D MP2RAGE acquisition was performed with the following parameters: TR/TI1/TI2/TE = 5000/700/2500/2.9 ms, voxel size = 0.7×0.7×0.7 mm (acquisition time ∼10 minutes).

At Institution B, 20 patients with early relapsing-remitting MS (RRMS) (75% female, 35±7 (mean ± standard deviation) years old, age range 21–46 years) with EDSS scores ranging from 0 to 4 (median 1.5), and disease duration < 5 years were imaged. A 7T research scanner (Siemens Healthcare, Erlangen, Germany) was used, and the protocol included 3D MP2RAGE (TR/TI1/TI2/TE = 6000/750/2350/2.92 ms, voxel size = 0.75×0.75×0.9 mm) and 3D T2*w multi-echo GRE (TR/TE1/TE2/TE3/TE4/TE5/TE6/TE7/TE8/TE9 = 45/4.59/9.18/13.77/18.35/22.94/27.53/32.12/36.71/41.3 ms, voxel size = 0.75×0.75×0.9 mm). For both datasets, MP2RAGE images were processed on the respective scanners with the Siemens research sequence package^24^ to obtain uniform denoised images and T1 maps. Throughout the manuscript, by MP2RAGE we always refer to its uniform denoised image^24^. The study was approved by the Institutional Review Board of both institutions, and all patients gave written informed consent prior to participation.

### Manual segmentation

In the 60 cases from institution A, the four uniform denoised (T1w) and T1 map MP2RAGE repetitions were co-registered and median T1w and T1map images were generated, as described previously^10^, median images are referred to as MP2RAGEx4 and MP2RAGE T1map x4 hereafter. CL were visually identified using MP2RAGEx4, MP2RAGE T1 map, T2*w GRE, and T2*w EPI images and delineated on MP2RAGEx4 independently by one neurologist (E.B.) and one neuroradiologist (J.M.) (both with several years of experience identifying CL), who subsequently reached consensus in a joint session. The experts classified the CL as leukocortical, intracortical, or subpial according to previously described criteria^1^. Cortical lesions were hypointense on MP2RAGE images and/or hyperintense on T2*w images and were seen on at least two consecutive axial slices. All lesions were manually segmented after consensus agreement using the image analysis software Display (http://www.bic.mni.mcgill.ca/software/Display/Display.html). In total, 2247 CL (21.0 median lesions/case, IQR=54) were segmented, of which 36% were leukocortical, 7% were intracortical, and 57% were subpial. This also includes 192 CL (37/8/147 leukocortical, intracortical, and subpial, respectively) that were added after a retrospective analysis by an expert of the “false positives” generated by the CNN in our previous study^24^. The intraclass correlation coefficient between the two raters was 0.91 (95% CI 0.85-0.94) for total CL, 0.91 (95% CI 0.85-0.94) for subpial lesions, and 0.91 (95% CI 0.85-0.94) for leukocortical lesions. Fig. 1 shows an example of each lesion type. A WML segmentation was obtained with a semi-automated method^26^.

**Figure 1.**
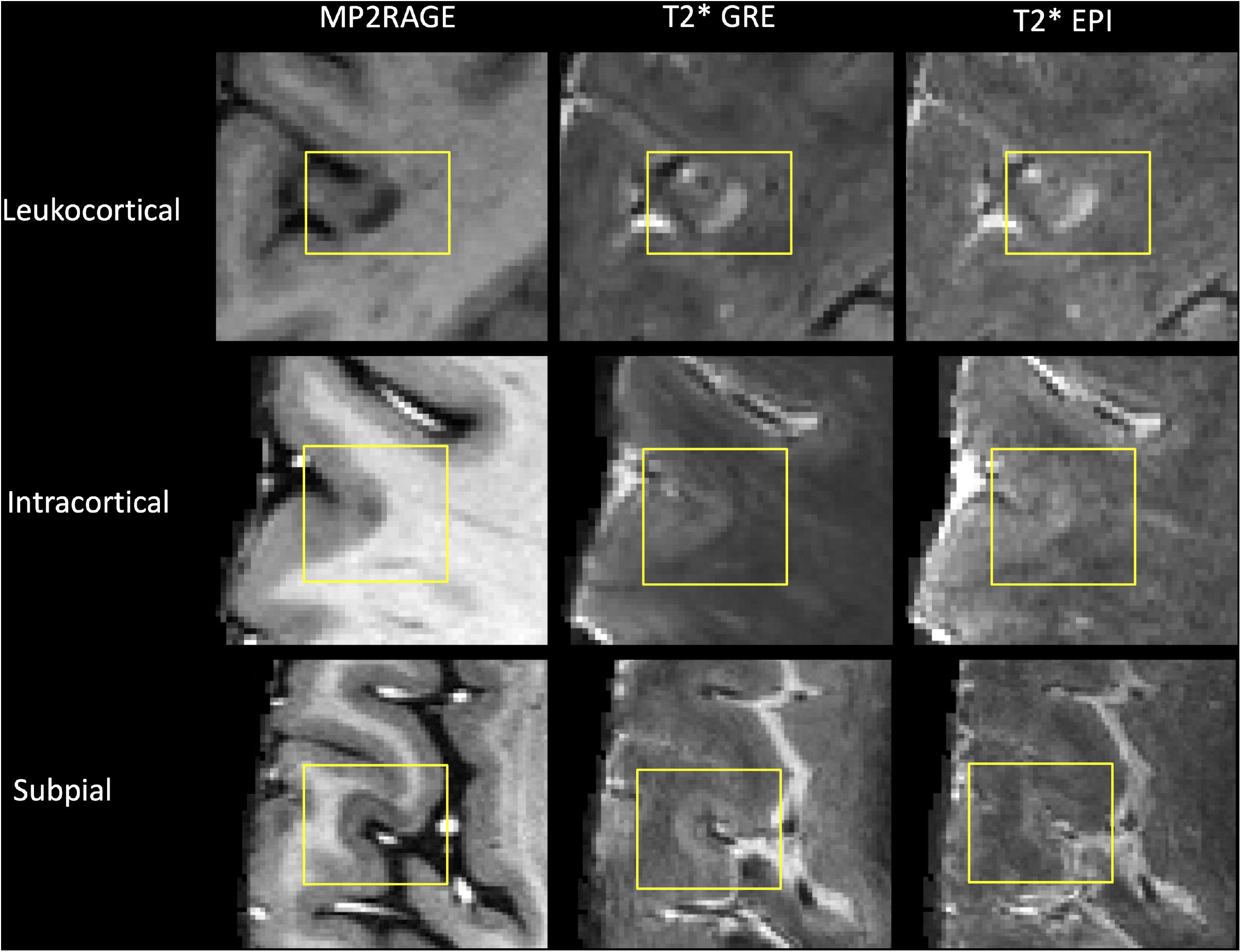
Examples of the three CL types identified in dataset A shown in the three MRI images considered.

In the cases from institution B, the manual segmentations were performed by consensus between 1 radiologist (A.T., 6 years of experience) and 1 neurologist with expertise in MS and neuroimaging (C.G., 13 years of experience). The ITK-SNAP (http://www.itksnap.org/) tool was used for the annotations. 188 CL (1.0 median lesions/case, IQR=10) were identified and subsequently classified into three types, leukocortical (69%), intracortical (26%), and subpial (5%). For the analysis of this dataset, intracortical and subpial lesions were grouped together.

### Pre-processing

The images of each subject were linearly registered to the same space (MP2RAGEx4) using ANTs^27^ and subsequently skull-stripped using FSL-Brain Extraction Tool^28^. All images were then resampled to 0.5×0.5×0.5 mm using a bilinear interpolation for the MRI contrasts and a nearest neighbor interpolation for the lesion masks. Finally, all non-zero voxels were normalized with mean 0 and standard deviation 1.

Examining the lesion masks, approximately 80 CL appeared to be outside of the T2* contrasts’ FOV and were therefore excluded in the analysis that included these contrasts.

### Convolutional neural network

CLAIMS relies on our previously proposed CNN architecture^23^ with several targeted modifications that boosted its performance. In particular, it is inspired by the 3D U-Net^28^, but we used four resolution levels instead of three, each one with an increasing number of features: 16, 32, 64, and 128, respectively. Compared to our previous work, the tissue segmentation output branch was removed, and bigger patches of size 96×96×96 voxels were provided as input to the network (see a scheme in Figure 2). As output, two separate labels are provided, one representing leukocortical lesions and another one representing intracortical and subpial lesions.

**Figure 2.**
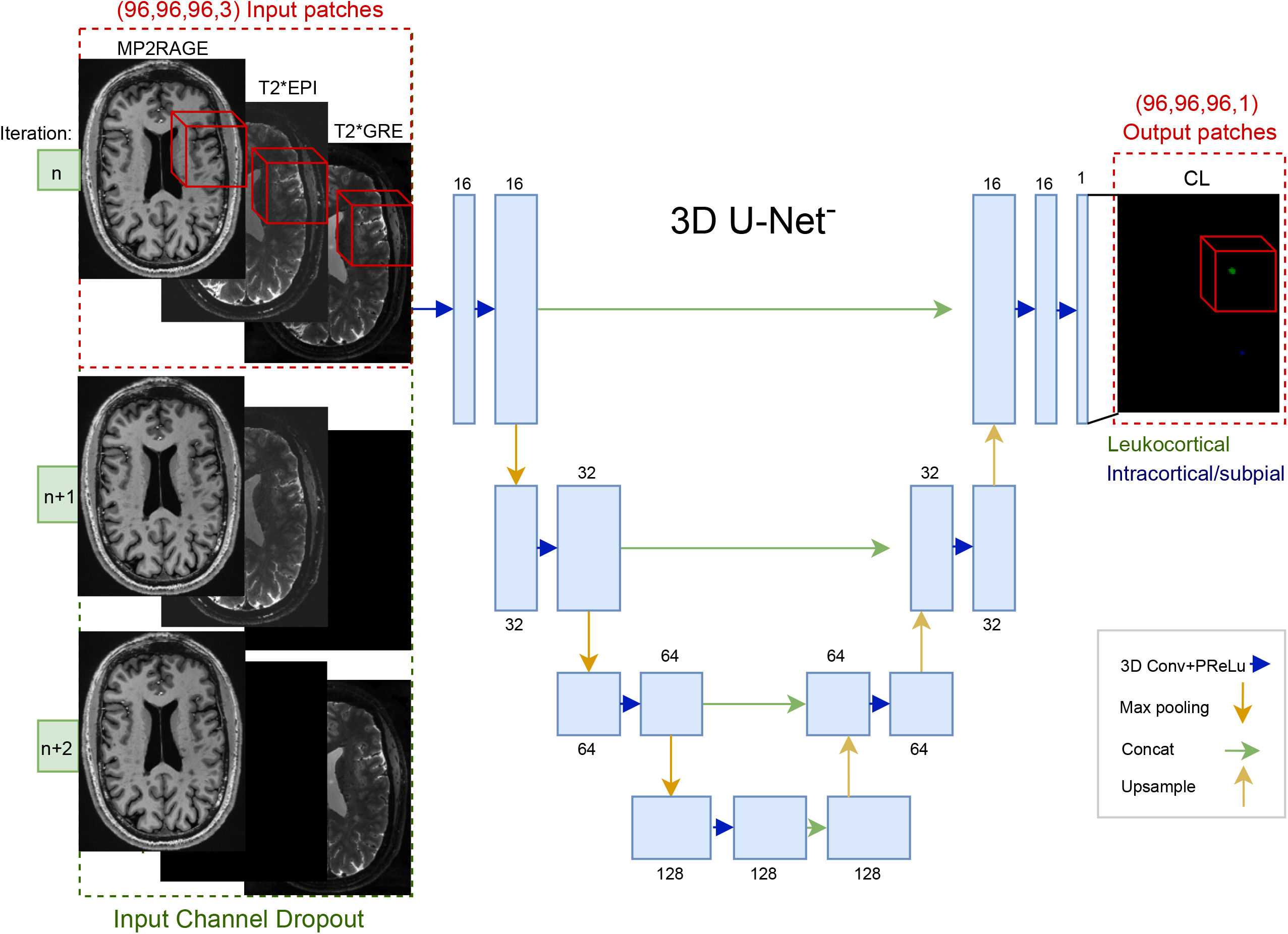
Scheme of the proposed multi-contrast CNN architecture inspired by the 3D U-Net. The CNN takes as input 3D patches of size 96×96×96 of the different MRI contrasts (red frame) and provides as output a CL detection and classification into two classes. Input channel dropout is applied when the two T2* contrasts are considered (green frame).

Instead of cross-entropy loss, used in our previous work^23^, we trained the network with focal loss (γ=2), as it has recently been shown to outperform cross-entropy for a large variety of tasks^29^. All WML voxels from the semi-automatically obtained WML masks were considered with a weight of 0 during training. Adam optimizer was used, and each model was trained for 400 epochs with an initial learning rate of 0.5e−3.

### Data augmentation

Extensive data augmentation was applied to reduce the risk of overfitting. Input sequence dropout was applied as in our previous study^24^, meaning that at each training iteration with multiple contrasts, one of the two T2* images is randomly dropped (eg. multiplied by zero). Moreover, random rotations of up to 180 degrees in the three planes and random flipping of the axis were applied. Additionally, random intensity shift (with an offset of 0.1) and affine transformations (random rotations of up to 15 degrees and random scaling of up to 10% of the image size) were performed.

The training was done on an NVIDIA RTX3090 for 400 epochs and took approximately 44 hours. Testing takes approximately 2 minutes for each single case using the same machine. The code has been implemented in MONAI^31^ running on top of Pytorch and is publicly available on our research website^1^.

### Experiments

Several experiments and ablation studies were performed in order to compare the different MRI contrasts and performance of CLAIMS on separate datasets.

- First, over the 60 cases of Institution A, a 6-fold cross-validation was done to compare the single and multi-image inputs (50 cases for training and 10 for testing in each fold). An internal validation was performed in each fold with 5 randomly drawn subjects. The following image combinations were considered as input: MP2RAGEx4 + T2* GRE + T2* EPI, MP2RAGEx4 + T2* GRE, MP2RAGEx4 + T2* EPI, MP2RAGEx4 alone, T2* GRE alone, and T2* EPI alone. The lesion count of the best performing model (MP2RAGEx4) was compared with ground truth’s lesion count. Moreover, we assessed the Spearman correlation between four disability measures and both automated and manual lesion counts.
- Second, an experiment was conducted to evaluate the performance of CLAIMS on MP2RAGE images obtained with a single acquisition and slightly bigger voxel size (acquisition time of approximately 10 minutes), which would be closer to a clinical scenario compared to the 0.5mm MP2RAGEx4 (with an acquisition time of about 40 min and therefore intended for research purposes). For this purpose, a model was trained either with the 0.5 mm MP2RAGEx4, alternatively with the MP2RAGE 0.7 mm isotropic images (see Fig. 3 for a visual example of the differences between these two images), and in a third approach by randomly providing at each iteration either a 0.5mm MP2RAGEx4 or a 0.7mm single acquisition MP2RAGE to the network. The resulting three models were then all tested on the 0.7mm single acquisition MP2RAGE images. Importantly, averaging four acquisitions increases the signal-to-noise ratio (SNR) by a factor of 2, whereas going from a voxel size of 0.5 to 0.7 increases the SNR by a factor of 2.7^32^. Therefore, 0.7mm single acquisition MP2RAGE has a slightly higher SNR than the 0.5mm MP2RAGEx4 (see Fig. 3).
- Third, training was performed with all cases of institution A (using both MP2RAGE 0.7mm single acquisition and 0.5mm MP2RAGEx4, but no T2* images) and then tested 14 cases of institution B. Further, a domain adaptation of this model with 6 different cases of institution B was also done, consisting of re-training all CNN’s layers starting with the previous weights. In this case, the fine-tuning lasted 50 epochs and had an initial learning rate of 0.5e-4. The results were compared with the previous state-of-the-art method using 14 subjects from the same dataset^23^.

### Evaluation and statistical analysis

Common detection metrics such as the lesion-wise true positive rate (LTPR) and lesion-wise false positive rate (LFPR) were considered for the evaluation on a lesion-wise level. Due to the extremely small size of CL (starting from 1 μL in our datasets), each lesion labeled by the expert is considered detected if it overlaps by at least one voxel with the automatically generated mask. The median detection rate and false positive rate are considered on a patient-wise level as well. The Dice coefficient (DSC) and volume difference (VD) were computed to quantify the accuracy of lesion delineation and volumetric segmentation for the subjects with at least one CL. The classification accuracy between the two types of CL considered (leukocortical vs intracortical/subpial) was assessed as the percentage of correctly classified CL. For all cases from institution A, a minimum lesion size of 1 μL was considered for both the automated and manual mask when computing the metrics, whereas for the subjects of institution B, we evaluated different minimum lesion volumes (1, 3, 6, 9, and 15 μL). For both datasets, the segmented connected components that overlap with WML are not considered as false positives, as the difference between leukocortical and juxtacortical lesions is subtle.

**Figure 3.**
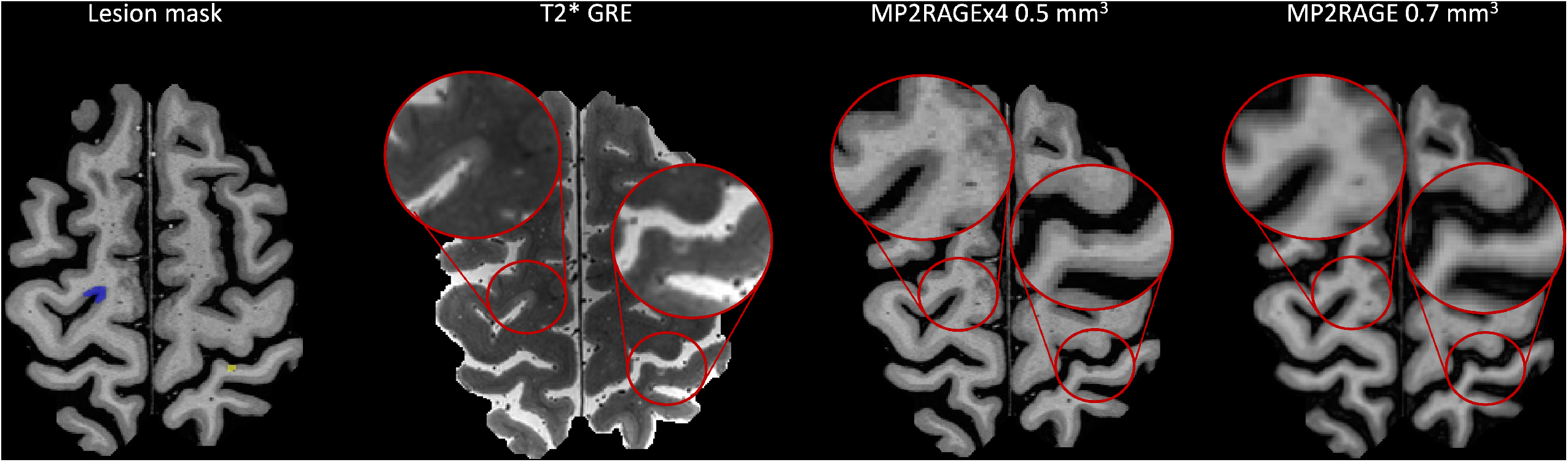
Examples of CL identified by the experts in the MP2RAGEx4 0.5mm and T2* GRE, and retrospectively seen in the MP2RAGE 0.7mm. A subpial lesion is marked by a blue mask, and an intracortical lesion is marked by a yellow mask.

The Wilcoxon signed-rank test was used to compare the metrics on a patient-wise level. The Bonferroni correction was used to adjust significance for multiple comparisons. Differences are considered significant at p-value < 0.05.

In order to verify the correlation between CL number and disability measures, the Spearman correlation coefficient (□) was computed. Cohen’s kappa coefficient (k)^33^ was evaluated to verify the agreement between the CL delineated by the experts and the CL detected by CLAIMS. Lin’s concordance coefficient (CCC)^34^ was computed to assess the level of correlation between the ground truth and CLAIMS’ lesion count.

## Results

### Single vs multi-image models

To assess the contribution of each MRI contrast, we performed the following ablation study. Three networks were trained, each with a single MRI contrast as input (MP2RAGEx4, T2*GRE, and T2*EPI), and then compared to two models receiving a bimodal input (MP2RAGEx4 and either T2*GRE or T2*EPI) and a model that received all three images (Figure 4, Table 1). In terms of lesion detection on a lesion-wise level, the three images model and the MP2RAGEx4 model achieved similar results, with a LTPR of 82% and 83% for leukocortical lesions, 49% and 53% for intracortical lesions, and 70% and 69% for subpial lesions. Both of them had a LFPR of 30%. They considerably outperformed the bimodal models and the single input models trained with T2*GRE and T2*EPI (LTPR of 34% and 30%, and LFPR of 45% and 49%, respectively). Similar performance was observed in terms of lesion delineation on a patient-wise level, with the 0.5mm MP2RAGEx4 achieving the best metrics (0.49/0.80 Dice coefficient and volume difference vs 0.18/4.51 for the T2*GRE and 0.16/4.57 for the T2*EPI). Finally, a moderately high CL subtype classification accuracy of over 80% was observed for all four models.

**Table 1.**
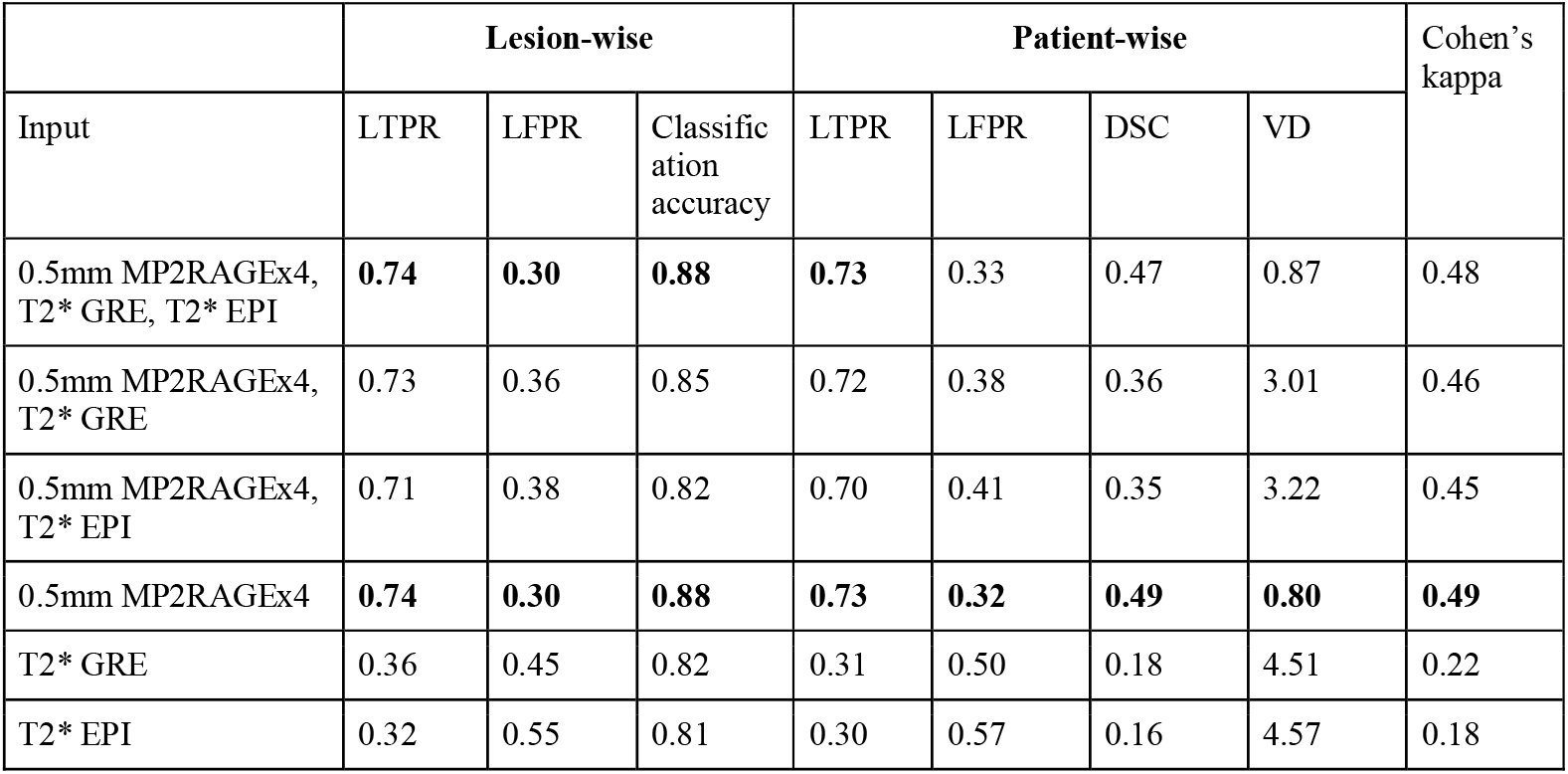
Median metrics and Cohen’s kappa coefficient for the different input contrasts obtained with a 6-folds cross-validation over the 60 cases of institution A. LTPR, LFPR and classification accuracy are computed on a lesion-level, whereas LTPR, LFPR, DSC and VD are considered on a patient-wise level. In bold the best result for each metric.

|  | Lesion-wise |  |  | Patient-wise |  |  |  | Cohen's kappa |
| --- | --- | --- | --- | --- | --- | --- | --- | --- |
| Input | LTPR | LFPR | Classification accuracy | LTPR | LFPR | DSC | VD |  |
| 0.5mm MP2RAGEx4, T2* GRE, T2* EPI | **0.74** | **0.30** | **0.88** | **0.73** | 0.33 | 0.47 | 0.87 | 0.48 |
| 0.5mm MP2RAGEx4, T2* GRE | 0.73 | 0.36 | 0.85 | 0.72 | 0.38 | 0.36 | 3.01 | 0.46 |
| 0.5mm MP2RAGEx4, T2* EPI | 0.71 | 0.38 | 0.82 | 0.70 | 0.41 | 0.35 | 3.22 | 0.45 |
| 0.5mm MP2RAGEx4 | **0.74** | **0.30** | **0.88** | **0.73** | **0.32** | **0.49** | **0.80** | **0.49** |
| T2* GRE | 0.36 | 0.45 | 0.82 | 0.31 | 0.50 | 0.18 | 4.51 | 0.22 |
| T2* EPI | 0.32 | 0.55 | 0.81 | 0.30 | 0.57 | 0.16 | 4.57 | 0.18 |

**Figure 4.**
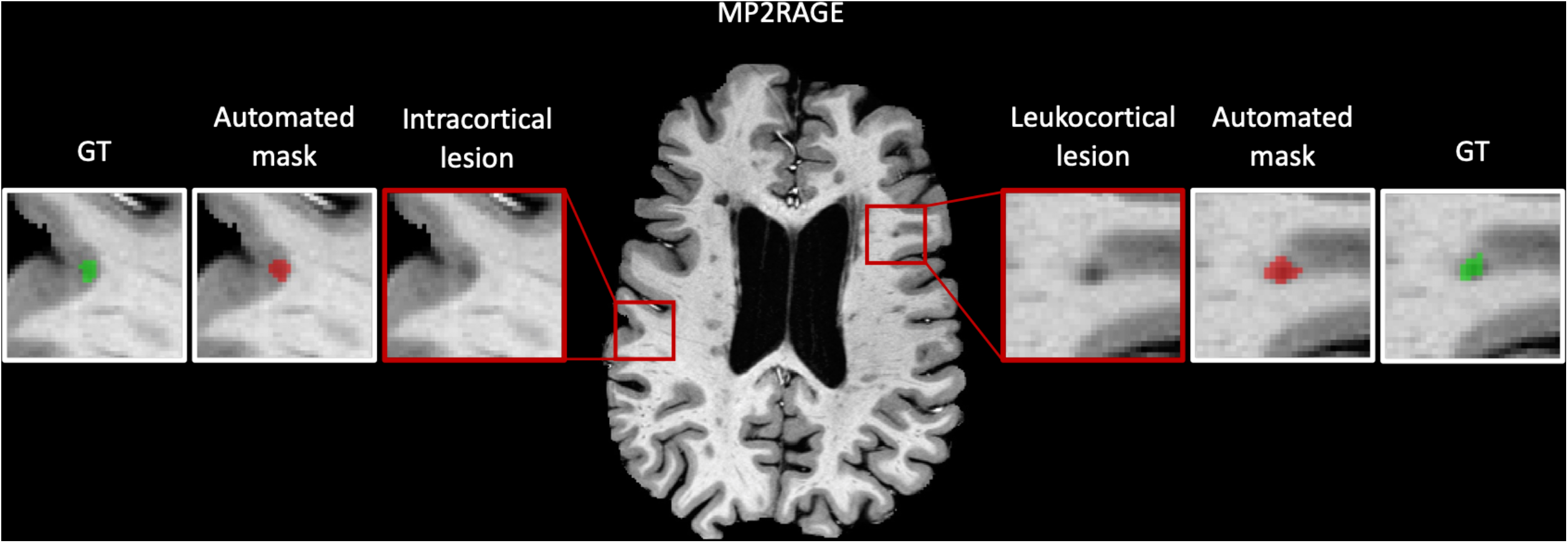
Visual results showing CL detection with the single input 0.5mm MP2RAGEx4 model. Left: an intracortical lesion manually segmented (green) that was correctly detected by the automated method (red). A similar example for a leukocortical lesion. GT=ground truth.

The patient-wise analysis in Fig. 5 shows the violin plots for the LTPR and LFPR for the single and three-input models. The single input 0.5mm MP2RAGEx4 model and the three images model achieved results with no significant statistical differences (p>0.05), and both significantly outperformed the single input models trained with T2*GRE and T2*EPI (p<0.001). The single input 0.5mm MP2RAGEx4 mode reached a Cohen’s kappa coefficient of 0.49 when compared with the manually annotated CL. Analyzing as well the correlation between the manual and automated lesion counts, Lin’s concordance coefficient reached 0.91 (see Fig. 6), indicating substantial agreement. Moreover, we computed the Spearman coefficient to compare the correlation between four disability measures (EDSS, 9-HPT, 25TW, SMDT) and both the ground truth and CLAIMS lesion counts. The manual and CLAIMS lesion counts correlated similarly with each of the disability measures, presented in Table 3. Comparable results were observed considering individual types of cortical lesions (Suppl. Table 2). The Bland-Altman plot was computed considering the volumetric differences between manual and automated CL segmentation (see Fig. 7). No particular bias was observed for CLAIMS segmentations.

**Table 2.** Comparison of lesion detection rate on a patient-wise level for the different models. NS: non-significant; *: p<0.05; **: p<0.01:***: p<0.001.

|  | MP2RAGEx4 |  | T2*GRE |  | T2*EPI |  | 3 contrasts |  |
| --- | --- | --- | --- | --- | --- | --- | --- | --- |
|  |  | Per patient |  | Per patient |  | Per Patient |  | Per Patient |
|  | Detected (%) | Median (range, IQR) | Detected (%) | Median (range, IQR) | Detected (%) | Median (range, IQR) | Detected (%) | Median (range, IQR) |
| All | 1649 (74%) | 14 (0-163, 36) | 748 (36%) | 6 (0-141,10) | 692 (32%) | 5 (0-128,11) | 1648 (74%) | 14 (0-151,31) |
| Leukocortical | 672 (83%) | 5 (0-54, 13) | 268 (63%) | 4 (0-39, 12) | 254 (59%) | 4 (0-36, 11) | 656 (82%) | 6 (0-48, 13) |
| Intracortical | 83 (53%) | 1 (0-17, 2) | 25 (15%) | 0 (0-5, 1) | 19 (12%) | 0 (0-2, 0) | 76 (49%) | 1 (0-17, 2) |
| Subpial | 894 (69%) | 6 (0-96, 8) | 455 (48%) | 4 (0-92, 6) | 421 (41%) | 5 (0-81, 4) | 916 (70%) | 7 (0-98, 9) |

Table 2. Comparison of lesion detection rate on a patient-wise level for the different models.
|  | Significance |  |  |  |  |  |
| --- | --- | --- | --- | --- | --- | --- |
|  | MP2RAGEx4 vs T2*GRE | MP2RAGEx4 vs T2*EPI | MP2RAGEx4 vs 3 contrasts | 3 contrasts vs T2*EPI | 3 contrasts vs T2*GRE | T2*GRE vs T2*EPI |
| All | *** | *** | N.S. | *** | *** | N.S. |
| Leukocortical | *** | *** | N.S. | *** | *** | N.S. |
| Intracortical | *** | *** | N.S. | *** | *** | ** |
| Subpial | *** | *** | N.S. | *** | *** | * |

**Table 3:** Spearman correlation coefficient ***ρ*** (and its relative p-value) computed between four disability measures and the manual and automated CL count. Both counts show a moderate correlation for all four measures.

|  | CLAIMS CL number |  | Ground truth CL number |  |
| --- | --- | --- | --- | --- |
| | $\rho$ | p-value | $\rho$ | p-value |
| Ground truth<br>CL number | 0.86 | <0.0001 | - | - |
| EDSS | 0.45 | 0.0003 | 0.43 | 0.0008 |
| 25TW | 0.42 | 0.0008 | 0.43 | 0.0008 |
| 9HPT | 0.50 | <0.0001 | 0.40 | 0.0014 |
| SDMT | -0.58 | <0.0001 | -0.53 | <0.0001 |

**Figure 5.**
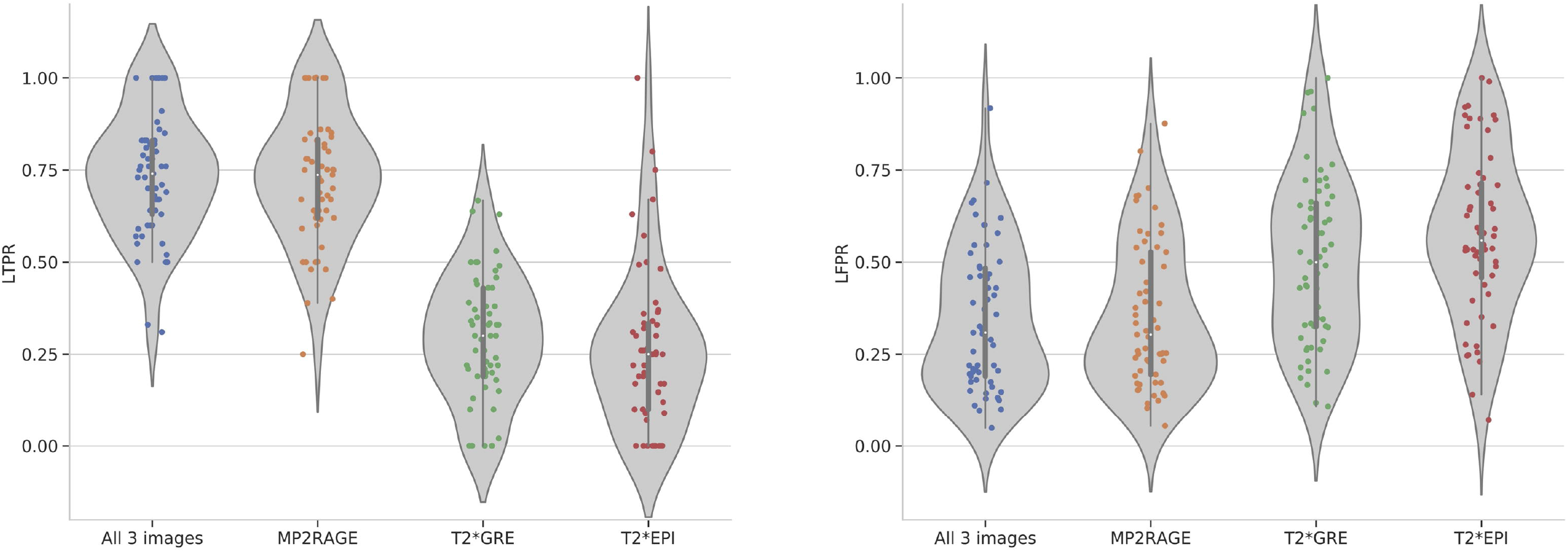
Violin plots of the LTPR and LFPR for different input models evaluated with a 6-fold cross-validation over the 60 subjects of institution A. Each dot represents a subject.

**Figure 6.**
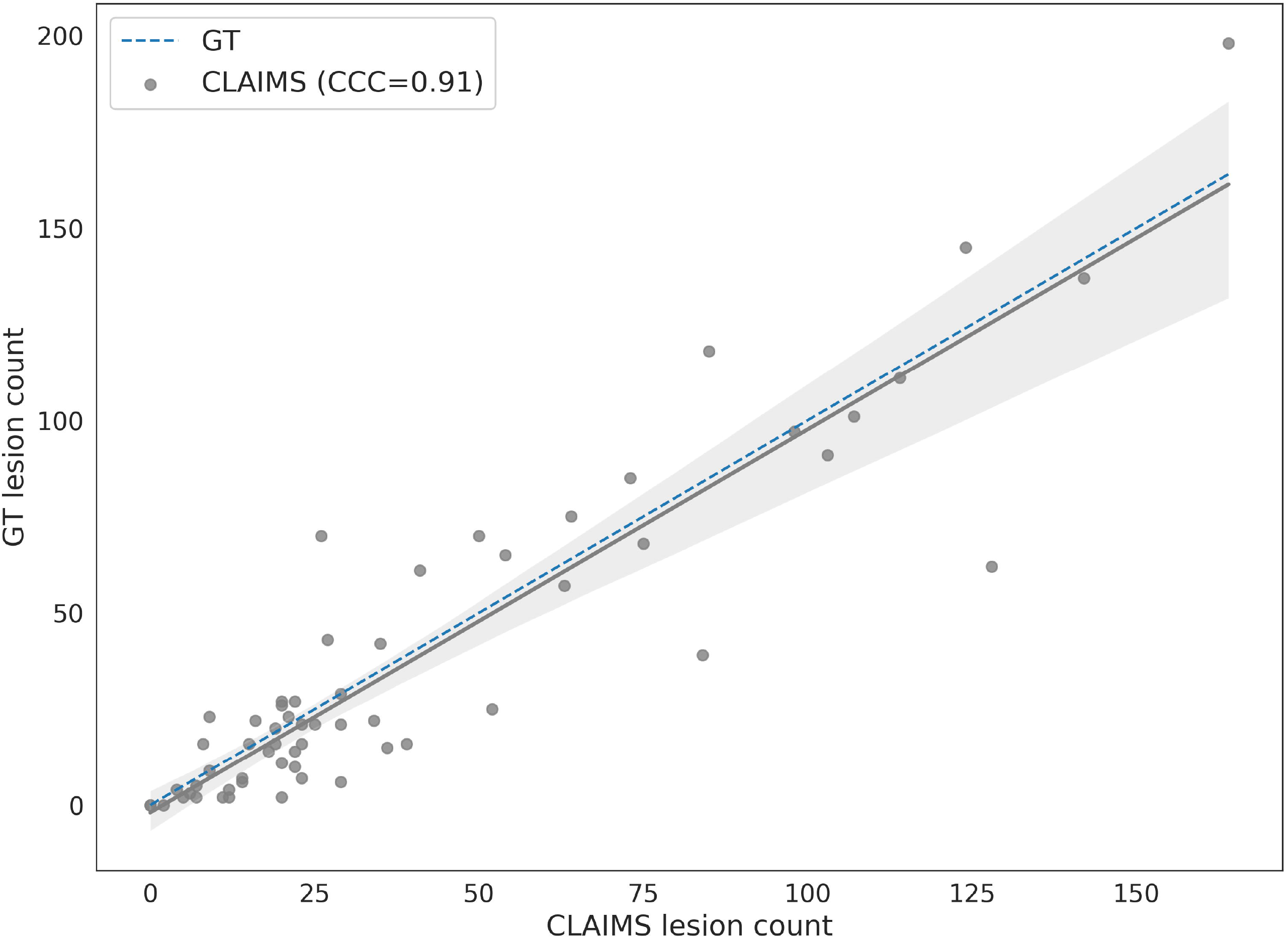
Correlation between the manual CL count and the one provided automatically by CLAIMS (best model trained with MP2RAGEx4). The solid lines show the linear regression model between the two measures along with a confidence interval at 95%. The dashed lines indicate the expected lesion count estimates. Lin’s concordance correlation coefficient between manual and automatic lesion count is reported in the legend (CCC = 0.91) and shows a substantial agreement.

**Figure 7.**
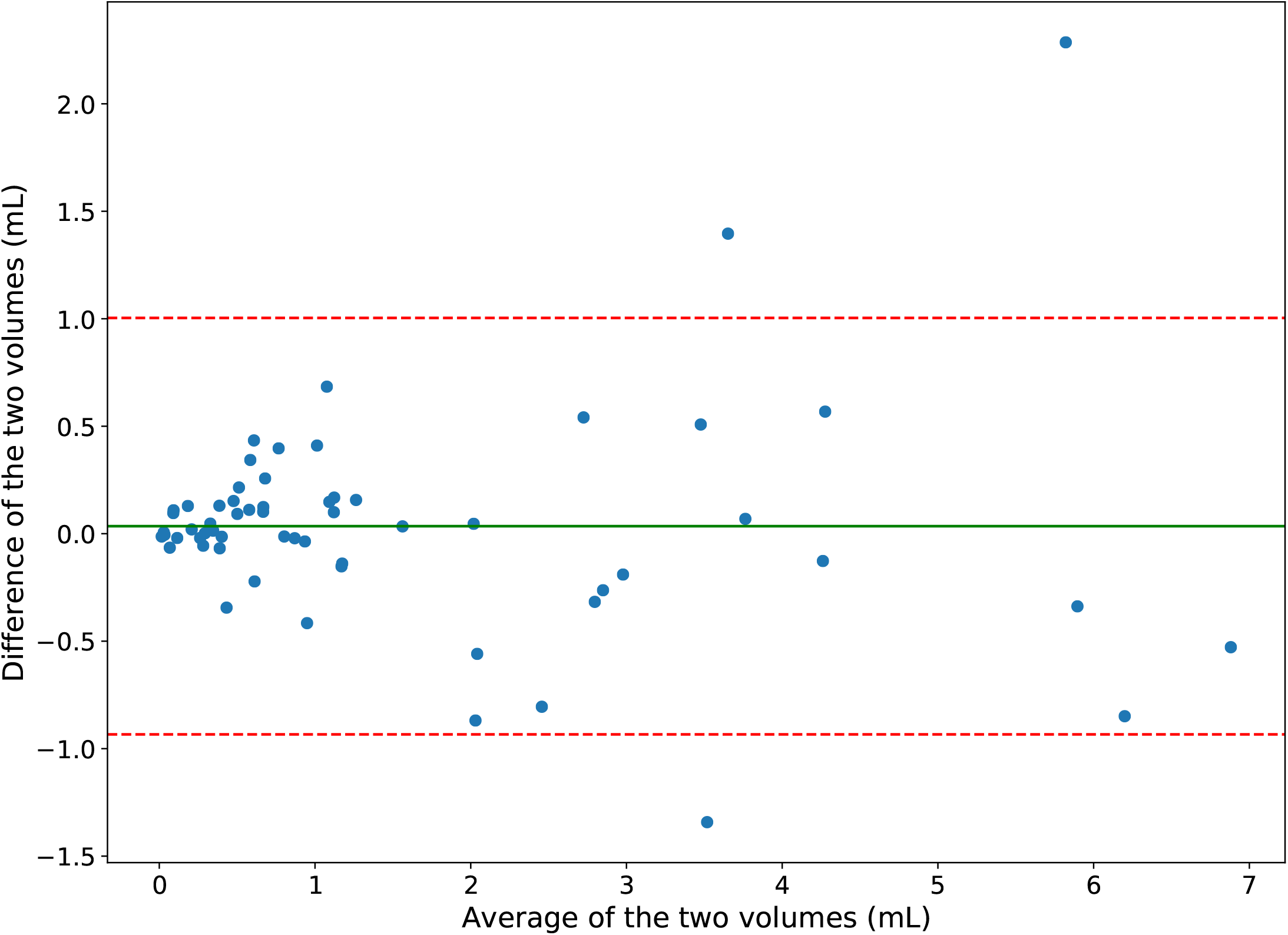
Bland-Altman plot (reference - prediction) of the manually and automatically segmented CL volumes. The solid green line shows the mean difference, whereas the dotted red lines the ±1.96 SD limits of the mean difference.

Table 2 and Fig. 8 show the detection rate for the three different CL types on a lesion-wise level. As observed in our previous work^24^, intracortical lesions remain the most challenging, with a detection rate of 53% in the best scenario (three image model). On the other hand, leukocortical and subpial lesions have a high detection rate of over 80 and 70%, respectively, for both the MP2RAGEx4 and the three images model. Similar behavior is observed between the three CL types for all models with a significant drop, however, in the detection of intracortical lesions for the T2*GRE and T2*EPI models (13% and 7%, respectively).

**Figure 8.**
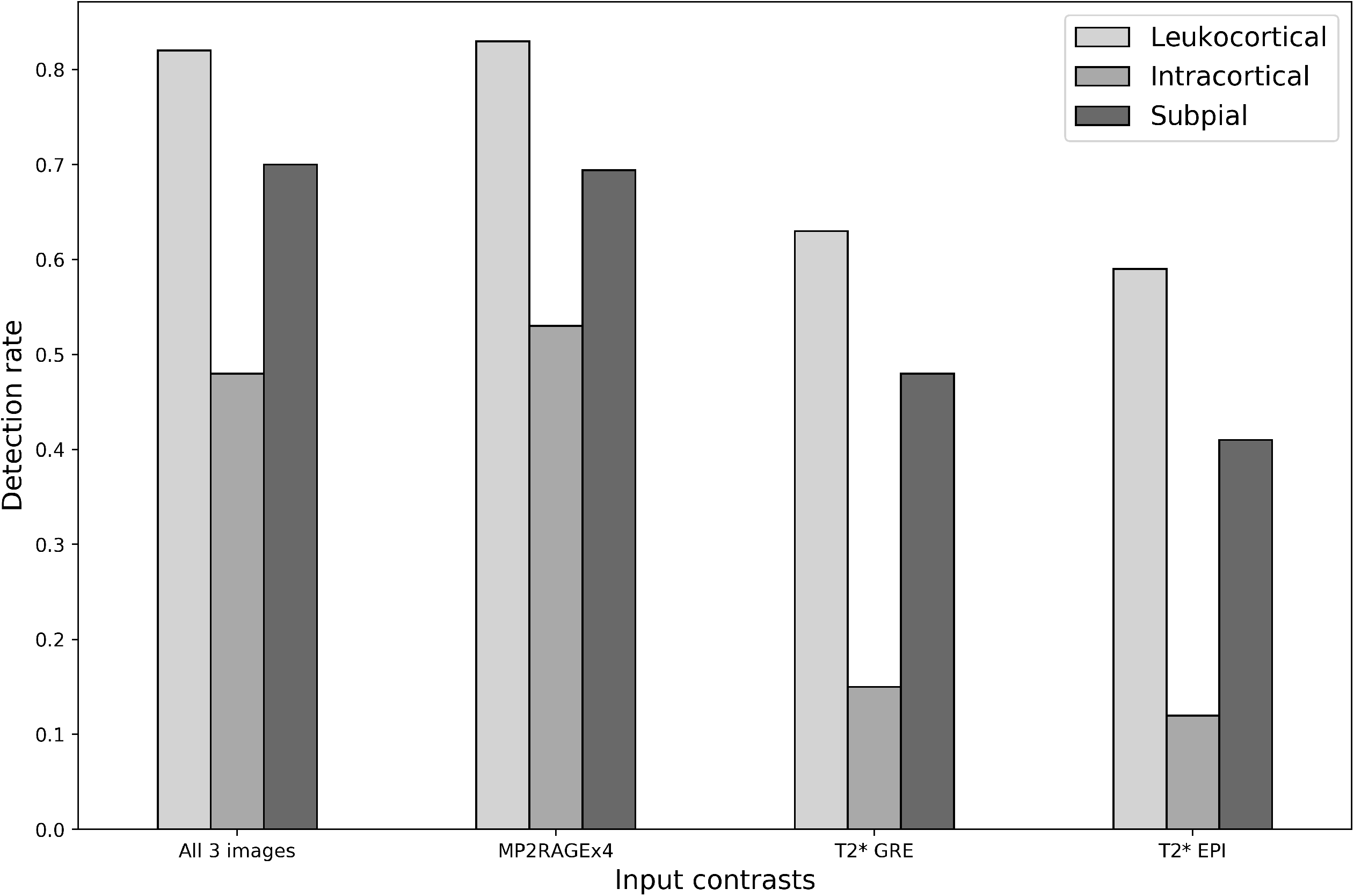
Lesion-wise CL detection rate for the three different CL lesion types considered over the 60 subjects of institution A.

### Evaluating CLAIMS on standard MP2RAGE images

In order to evaluate CLAIMS on more commonly acquired single acquisition 0.7mm MP2RAGE images, three models were trained with the following inputs: (1) MP2RAGE 0.7mm, (2) MP2RAGEx4 0.5mm, (3) alternating MP2RAGEx4 0.5mm and 0.7mm at each training iteration. All models were then tested on the single acquisition 0.7mm MP2RAGE. For any given subject, care was taken to ensure that all images were either in the training or testing set and not split across them. Table 4 reports the metrics obtained when evaluating these models on the MP2RAGE 0.7mm images with a 6-fold cross-validation. All model results were evaluated with respect to the ground truth determined using MP2RAGEx4, T2*w GRE, and T2*w EPI images. First, compared to the previous results for the 0.5mm MP2RAGEx4 single input model (see Table 1), we observed that training and testing on single acquisition 0.7mm MP2RAGE causes a drop in the detection rate from 74 to 53%. Second, between the three models, the one using both 0.5 and 0.7mm images during training achieves the best metrics with LTPR of 53% and LFPR of 33%. Finally, the worst-performing model is the one trained only with MP2RAGEx4, having LTPR of 35% and LFPR of 41%.

**Table 4.** Metrics obtained for models trained with different inputs (listed in the first column) and tested on the MP2RAGE 0.7mm images. In bold is shown the best result for each metric.

|  | Lesion-wise |  |  | Patient-wise |  |
| --- | --- | --- | --- | --- | --- |
| Training images | LTPR | LFPR | Classification accuracy | Dice | VD |
| MP2RAGE 0.7mm | 0.52 | 0.39 | **0.85** | 0.25 | 1.10 |
| MP2RAGEx4 0.5mm | 0.35 | 0.41 | 0.81 | 0.16 | 1.29 |
| MP2RAGEx4 0.5 and<br>MP2RAGE 0.7mm | **0.53** | **0.33** | **0.85** | **0.29** | **1.06** |

### Independent test set

In our last study, an MP2RAGE-only model was trained with all cases from institution A (mixing 0.5mm MP2RAGEx4 and 0.7mm MP2RAGE, as this was the top performing input in the previous experiment) and tested on the 14 subjects of Institution B, which include only 0.75×0.75×0.9mm MP2RAGE images and were used also to report results with MSLAST in a previous work^23^. Furthermore, we also propose a domain adapted version of CLAIMS (CLAIMS_DA), fine-tuning the same model (training all layers) on 6 additional cases from Institution B. Fig. 9 presents the lesion-wise true and false positive rates for CLAIMS, CLAIMS_DA, and MSLAST, considering different minimum lesion volumes, even much smaller than the ones proposed in the MSLAST paper (1, 3, and 9μL vs 6 and 15μL). CLAIMS outperforms MSLAST for all volume thresholds and CLAIMS_DA achieves even better results on a lesion-wise level. In particular, considering a minimum lesion volume of 6μL, it reaches a detection rate of 71% with a false positive rate of 29% (see Supplementary Material Table 3). The CL classification accuracy for CLAIMS and CLAIMS_DA was 81% and 84%, respectively. On a patient-wise level, no statistically significant differences were observed between the three models.

**Figure 9.**
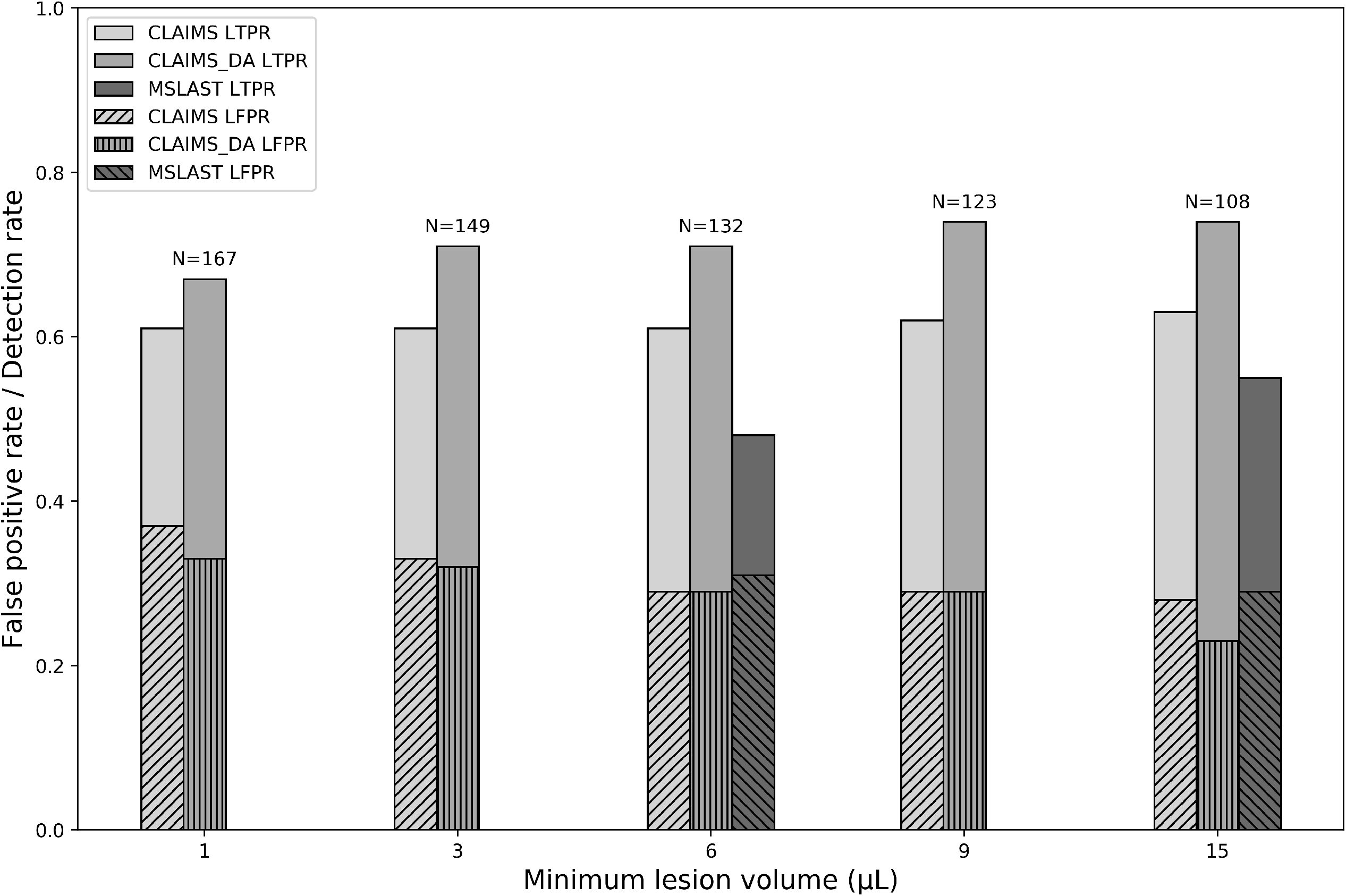
False positive and detection rate in a pure testing scenario on the Institution B dataset for CLAIMS, CLAIMS domain-adapted (CLAIMS_DA), and MSLAST^22^. Different minimum lesion volumes are considered. N refers to the number of CL in the ground truth for each minimum lesion volume.

## Discussion and Conclusion

In this work, we explored the value of different 7T MRI contrasts for the automated detection of CL. For this purpose, we trained and tested a novel U-net-based deep learning method we call CLAIMS. We analyzed three MRI contrasts as input (MP2RAGE, T2*-w GRE, and T2*-w EPI), different image resolutions, and CLAIMS’s generalizability on an external testing dataset. CLAIMS’ automated CL count was then compared to the experts’ CL number, and its correlation with four disability measures was examined. Furthermore, we compared our results with the only previous automated CL segmentation method at 7T in the literature^23^. Lastly, we reported the results obtained considering different minimum lesion volumes and analyzing the three CL subtypes.

The results of our ablation study with different input contrasts fed to the model show that the MP2RAGE contrast alone is sufficient to achieve the best performance both on a lesion and patient-wise level. Contrary to a past study regarding the manual segmentation of CL^6^, the addition of two T2*-weighted images did not contribute significantly to improving the automated CL detection. Moreover, the single input models trained with the T2*-weighted contrasts performed poorly compared to the MP2RAGE one. This is in line with a previous study where it was shown that the MP2RAGE increases the visual detection of all CL types compared to T2*-weighted imaging at 7T^10^. However, it is important to note that our ground truth was created considering all three MRI images, and some lesions might not be visible on the T2* contrasts alone. Moreover, even a lower resolution MP2RAGE outperformed the T2*-weighted models, arguably establishing the MP2RAGE as the preferred contrast by CLAIMS. Focusing on the different CL types, we observed that their detection rate varies considerably. As in our previous study^24^, intracortical lesions were once again the most challenging type, with a detection rate of only 53% for the best model. On the other hand, both leukocortical and subpial lesions were detected with a high sensitivity of over 80 and 70%, respectively. These values approach the inter-rater agreement of ∼85% in a previous study considering two raters^11^. In the prior study, Cohen’s kappa coefficient was 0.69, showing substantial agreement, whereas ours, computed between CLAIMS output and the manual masks, reached 0.49. However, we considered a dataset with twenty times more CL (2247 vs 103), and this could explain the higher variability. Similarly, the inter-rater reliability can also be estimated by computing Lin’s concordance correlation coefficient, which takes into account the correlation between lesion counts. Analyzing lesion counts from 10 sample scans and by two raters, Harrison et al. showed Lin’s coefficient of 0.54 for all CL^3^, meaning that the agreement was weak. On the contrary, our best model achieved a value of 0.91, showing a substantial correlation between the manual and automated CL count. Moreover, CLAIMS computes a single subject’s lesion map in approximately 2 minutes, whereas manual raters needed anywhere between 30 min and several hours depending on lesion burden to perform a similar task manually. This suggests that CLAIMS could be useful to support and speed up experts’ CL assessments. To further support this claim, we analyzed the correlation between four disability measures and the manual and automated CL numbers. Both CL counts correlated with the four measures (Spearman’s coefficient between 0.40 and 0.57) in a very similar way. To the best of our knowledge, this is the first time that an automated CL count has proven to correlate with disability scores.

Relying only on the MP2RAGE, we then evaluated CLAIMS on MP2RAGE images that are more feasible to acquire, obtained with a single acquisition of about 10 minutes (vs 40min for MP2RAGEx4) and with a slightly bigger voxel size. Specifically, we compared different models trained with either 0.5mm, 0.7mm, or both 0.5mm and 0.7mm. The 0.5mm isotropic images were obtained as an average of 4 acquisitions and had, therefore, an SNR twice as high compared to that of a single acquisition at the same resolution, but a lower SNR compared to the 0.7mm MP2RAGE. Importantly, the model trained with 0.5mm images and tested on 0.7mm images (simply interpolated to 0.5mm) performed very poorly, showing the value of the voxel size for the detection of very small structures. It is important to keep in mind that the ground truth was labelled on the 0.5mm images, and this might also cause a partial drop in performance. However, when mixing both 0.5mm and 0.7mm images during training, the metrics improve considerably, even outperforming the model trained with 0.7mm images only. This indicates that CLAIMS can extract useful information during training from the smaller voxel size images and successfully use these at inference time. Overall, however, we notice that going from 0.5mm MP2RAGEx4 to single acquisition 0.7mm MP2RAGE causes a detection rate drop of about 20%, proving the importance of image quality and resolution even for automated methods.

Finally, we tested the performance of CLAIMS trained mixing both 0.5mm MP2RAGEx4 and 0.7mm MP2RAGE on a different dataset where MP2RAGE scans were acquired with a voxel size of 0.75×0.75×0.9mm. This is the same dataset on which the previous state-of-the-art method (MSLAST) was evaluated^23^, and therefore we could carry out a precise comparison including the same subjects. It is important to note that the dataset from Institution B includes subjects in the very early stages of the disease (disease duration < 5 years) who have a much lower lesion burden compared to the subjects of institute A (median lesion count per case of 1 vs 21). Moreover, the CL subtype distribution is different as well, with a majority of leukocortical lesions in dataset A, whereas intracortical lesions are prevalent in dataset B (see Supplementary Material, Suppl. Fig. 1). This could be partially explained by the lower voxel size and the presence of T2* images in dataset A, which allow higher visual detection of subpial lesions^6^. Nevertheless, CLAIMS proves to be robust and performs well in this multi-center scenario, with a lesion-wise detection rate of about 61% when different minimum lesion volumes are considered (1, 3, 6, 9, and 15μL). It outperforms MSLAST (both for minimum lesion volume of 6μL and 15μL), and it also classifies CL into two types (leukocortical and intracortical) with an accuracy of about 80%. When decreasing the minimum lesion volume considered, the detection rate remains stable, with only a marginal increase of false positives. For instance, if a minimum lesion volume of 3μL is considered, CLAIMS achieves a detection rate of 61% with a false positive rate of 33%. Moreover, when fine-tuning CLAIMS with six additional subjects belonging to the same dataset, the lesion-wise performance increased considerably, reaching a detection rate of 71% with a false positive rate of 32%. This confirms the efficacy of domain adaptation for deep learning-based models applied to a different dataset^35^ and poses our proposed method as the state-of-art technique for CL detection with 7T MRI.

In this study, results were evaluated both on lesion and patient-wise levels. During training, our proposed CNN takes as input random 3D patches extracted from different patients. As the CNN does not see the field of view of the entire brain, a lesion-wise evaluation is crucial to determine its performance at testing time. At the same time, however, from a clinical research perspective, when evaluating the effects of CL on clinical outcomes, patient-wise data is the most relevant. Moreover, the lesion-wise detection rate might be extremely dependent on cases with a high lesion load while not reflecting the method’s performance on subjects with few lesions. For these reasons, both analyses were considered to provide a comprehensive evaluation.

A recent study has investigated the dependence between the training dataset size and the segmentation performance of a convolutional neural network^37^. Considering a 2D U-Net architecture, Narayana et al. concluded that 50 subjects are sufficient for an accurate MS lesion segmentation^37^. In our study, we designed a 3D CNN architecture instead, while, however, having a reduced number of feature maps and resolution levels compared to the original 2D U-Net architecture. Each training fold included 50 subjects. Thus, we believe that our sample size is adequate for the task, and this is supported also by the promising results obtained on the external testing dataset (Center B). Moreover, considering subjects from both Center A and B, we analyzed the largest cohort of MS patients for automated CL detection in the literature^38^.

Our study also presents some limitations. First, the two datasets considered were acquired with scanners from the same manufacturer and with similar acquisition protocols. Additional differences in the images could arise under more general conditions, potentially causing a drop in performance. In this case, a fine-tuning of the pre-trained model relying on a few annotated subjects could help overcome this issue and help regain performance. Second, the detection rate of intracortical lesions remains quite low, even though it improved by more than 10% compared to our previous study^24^. These are infrequently visible and extremely small CL, and perhaps a larger training dataset with an increased number of intracortical lesions could mitigate this issue. Of note, there was a difference in relative prevalence of intracortical vs subpial lesions in the two datasets, likely related to differences in lesion appearance between the two imaging protocols (superficial cortical involvement is often more apparent on T2*w images). By grouping intracortical and subpial lesions for the purposes of the model, we were able to achieve good performance and lesion classification on both datasets, however differentiating between intracortical and subpial lesions remains a difficult task, both for manual rating and automated methods. Third, in this work, we analyze a single CNN architecture that proved effective for CL detection in our previous study. It was out of the scope of this study to compare several different models and tweak their parameters to optimize the performance. We rather selected the state-of-the-art architecture and analyzed in detail its potential to tackle a clinically relevant problem such as the detection of CL. Fourth, although the MRI sequences used here are highly sensitive for CL compared to 3T techniques, their true sensitivity to CL is unknown and it is likely that some CLs are not well seen using these techniques. Thus, some of the false positive lesions detected by the method presented here may be true lesions, as shown in our previous study^24^.

Future work could include exploring more advanced deep learning architectures in order to better include the information coming from each single MRI contrast, particularly the T2*w images. Given time constraints in the clinical setting, use of fewer MP2RAGE acquisitions could be explored as well to determine how much each additional repetition contributes to improved lesion sensitivity. In addition, novel compressed sensing techniques^36^ could be exploited to shorten the MP2RAGE acquisition time to a clinically acceptable duration even with multiple repetitions. Moreover, the use of larger and additional datasets could further prove the generalizability of the proposed method. Finally, the use of the T1 map image type generated from MP2RAGE acquisitions could also be explored for the automated detection of CL, although in our experience most CL are similarly seen on both MP2RAGE uniform denoised and T1 map images.

In conclusion, we present CLAIMS, a DL framework for the detection and classification of CL with 7T MRI. When CLAIMS is trained only with MP2RAGE, it achieves state-of-the-art performance for all three CL types, and its CL count correlates with disability measures similarly to experts’ visual assessment. If fine-tuned, it adapts extremely well to a different dataset acquired in a different site. As 7T scanners from several manufactures are now being approved for clinical use, CLAIMS could eventually be useful to support clinical decisions, particularly in the field of diagnosis and differential diagnosis of MS patients.

## Data Availability

All data produced in the present study are available upon reasonable request to the authors.

## Abbreviations

MS: multiple sclerosis
GM: gray matter
WM: white matter
CL: cortical lesions
WML: white matter lesions
MRI: magnetic resonance imaging
3D: three-dimensional
T2* EPI: T2*-weighted segmented echo-planar imaging
T2* GRE: T2*w multi-echo GRE
FLAIR: 3D T2-weighted fluid-attenuated inversion recovery
MP2RAGE: magnetization-prepared 2 rapid acquisition gradient echoes
MPRAGE: magnetization-prepared rapid acquisition gradient echo
SNR: signal-to-noise ratio
CCC: Lin’s concordance correlation coefficient
k: Cohen’s kappa coefficient
CNN: convolutional neural network
CLAIMS: Cortical Lesion AI-based assessment in Multiple Sclerosis
EDSS: Expanded Disability Status Scale
9-HPT: 9-Hole Peg Test
25TW: 25-foot timed walk
SDMT: Symbol Digit Modalities Test
LTPR: lesion-wise true positive rate
LFPR: lesion-wise false positive rate
DSC: dice coefficient
VD: volume difference

## Acknowledgments

We thank members of the Translational Neuroradiology Section and the staff of the National Institute of Mental Health Functional MRI Facility for help with MRI acquisitions. We acknowledge access to the facilities and expertise of the CIBM Center for Biomedical Imaging, a Swiss research center of excellence founded and supported by Lausanne University Hospital (CHUV), University of Lausanne (UNIL), Ecole Polytechnique fedérale de Lausanne (EPFL), University of Geneva (UNIGE) and Geneva University Hospitals (HUG). We thank Bénédicte Marechal for the help with MSLAST and Thomas Yu for proofreading the manuscript.

## Footnotes

1 https://github.com/Medical-Image-Analysis-Laboratory

