## Supplementary material for "Multiple sclerosis cortical lesion detection with deep learning at ultra-high-field MRI"

|  | Institution A | Institution B |
| --- | --- | --- |
| Number of subjects | 60 | 20 |
| Females/Males | 38/22 | 15/5 |
| Age mean ± standard deviation [range] years | 49±11 [29–77] | 35±7 [21–46] |
| EDSS median [range] | 2.0 [0-7.5] | 1.5 [0-4.0] |
| Total number of CL | 2247 | 192 |
| Median number of CL per subject (IQR) | 21 (54) | 1 (10) |

Supp. Table 1. Metadata information of the two datasets considered.

|  | CLAIMS leukocortical lesion number | | Ground truth leukocortical lesion number | | CLAIMS intracortical/subpial lesion number | | Ground truth intracortical/subpial lesion number | |
| --- | --- | --- | --- | --- | --- | --- | --- | --- |
|  | 𝝆 | p-value | 𝝆 | p-value | 𝝆 | p-value | 𝝆 | p-value |
| EDSS | 0.46 | 0.0002 | 0.47 | 0.0002 | 0.37 | 0.0037 | 0.46 | 0.0003 |
| 25TW | 0.39 | 0.0021 | 0.39 | 0.0025 | 0.37 | 0.004 | 0.41 | 0.0011 |
| 9HPT | 0.43 | 0.0006 | 0.38 | 0.0025 | 0.40 | 0.0016 | 0.38 | 0.0028 |
| SDMT | -0.57 | <0.0001 | -0.47 | 0.0002 | -0.52 | <0.0001 | -0.48 | 0.0002 |

Supp. Table 2. Spearman correlation coefficient 𝝆 (and its relative p-value) computed between four disability measures and the manual and automated CL count, subdivided per CL type. Both counts show a moderate correlation for all four measures.

|  | Lesion-wise | | |
| --- | --- | --- | --- |
| Models | LTPR | LFPR | Classification Accuracy |
| CLAIMS | 0.61 | **0.29** | 0.81 |
| CLAIMS_DA | **0.71** | **0.29** | **0.84** |
| MSLAST | 0.48 | 0.31 | - |

Supp. Table 3. Lesion-wise metrics obtained with a minimum lesion volume of 6μL for CLAIMS, CLAIMS_DA, and MSLAST. In bold are the best results for each metric. No significant differences (p>0.05) were observed between the three models on a patient-wise level for both LTPR and LFPR. MSLAST did not classify CL in multiple types.

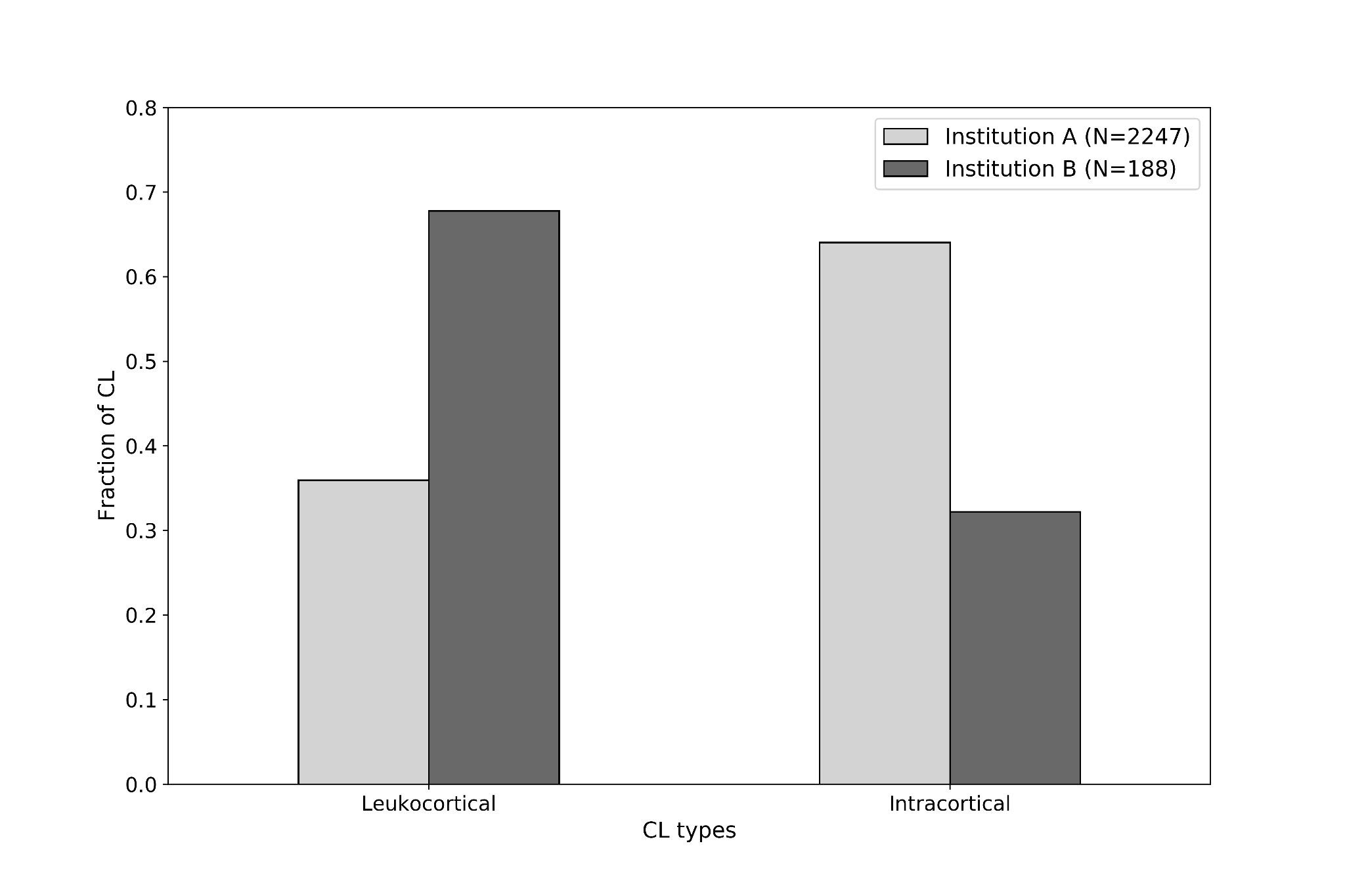

Supp. Figure 1. Fraction of leukocortical and intracortical/subpial CL in dataset A and B. N refers to the total number of CL in each dataset.
